# Diabetes Management Beliefs among Adults Diagnosed with Type 2 Diabetes in Iran: A Theory Informed Approach from a Theory of Planned Behavior Framework

**DOI:** 10.1101/2022.04.05.22273415

**Authors:** Mohammad Payam Ghaffari, Katherine M. White, Kourosh Djafarian, Susie Cartledge, Seyed Ali Keshavarz, Reza Daryabeygi-Khotbehsara, Sheikh Mohammad Shariful Islam

## Abstract

**Objective:** The current study was informed by the belief basis of Ajzen’s (1991) Theory of Planned Behavior (TPB) to identify the important behavioral (advantages and disadvantages), normative (important referents) and control (barriers and facilitators) beliefs associated with the key recommended prevention and management behaviors for adults in Iran diagnosed with Type 2 diabetes (T2D).

**Methods:** A cross-sectional study was conducted. A total of 115 adults diagnosed with T2D completed a questionnaire examining behavioral, normative and control beliefs and intention in relation to the three diabetes management behaviors including low fat food consumption, carbohydrate counting and physical activity. For each behavior, intention was considered as dependent variable; beliefs were independent variables. Analyses involved three multivariate one-way analysis of variance (MANOVAs).

**Results:** The findings for carbohydrate counting and physical activity suggested behavioral and control beliefs as differentiating high from low intenders to perform the behavior. For carbohydrate counting, behavioral beliefs such as weight control, improving one’s health, feeling good and controlling diabetic complications differed significantly between low and high intenders. For physical activity, feeling good, controlling blood sugar and tiredness were among behavioral beliefs differentiating low and high intenders. Medical advice from professionals and greater knowledge were identified as facilitators of carbohydrate counting. High costs were identified as a key barrier preventing individuals from engaging in physical activity. Spouse was the single significant referent influencing carbohydrate counting.

**Conclusions & Implications:** Identifying the underlying beliefs of key diabetes management behaviors can assist in the design of tailored educational interventions for individuals with T2D.

## Introduction

Diabetes is a fast-growing chronic disease with Type 2 diabetes (T2D) the most common type usually affecting older adults, although increased prevalence is now seen in young adults and even children (1). T2D can be initially controlled by healthy eating and physical activity, even though over time most people with diabetes will need medication and insulin (2). According to the International Diabetes Federation (IDF), the global prevalence of diabetes in the adult population (20 to 79 years) is estimated at 10.5% equivalent to 537 million in 2021 which is projected to reach 12.3% equivalent to 783 million in 2045 (3). In the same report, age-adjusted prevalence of diabetes in Iran is estimated to be 9.1% equivalent to 5.5 million in 2021 (3). People diagnosed with diabetes are prone to debilitating and threatening health outcomes. Diabetes outcomes in the long-term progressively cause specific complications including retinopathy, nephropathy, neuropathy and cardiovascular effects (3, 4). Maintenance of blood glucose, cholesterol and blood pressure near normal levels can delay or prevent diabetes complications (3).

Control of these key modifiable health parameters can be assisted by diabetes education, which is the cornerstone of diabetes control. The ADA (American Diabetes Association) recommends lower saturated fat intake in individuals diagnosed with diabetes and considers carbohydrate monitoring using carbohydrate counting as a key strategy in the management of blood glucose (5). Basic carbohydrate counting is suggested as essential, even for T2D patients who don’t use oral medication or insulin, to better control their disease (6). Physical activity also is a key component in T2D management (7, 8).

Many psychological theories have been proposed to understand the factors that may affect people’s health related decisions such as the Health Belief Model [HBM] (9), Theory of Reasoned Action [TRA] (10), Theory of Planned Behavior [TPB] (11), Social Cognitive Theory [SCT] (12) and Trans-Theoretical Model [TTM] (13). Among these, the Theory of Planned Behavior (TPB) (11), has been commonly used to examine people’s intention to perform a wide variety of health prevention and management behaviors. According to the TPB, a person’s behavior is a function of intention to engage in the behavior. In turn, intention is directly determined by a person’s attitude (positive or negative evaluation of the behavior), subjective norm (perceived pressure to perform or not perform a behavior), and perceived behavioral control (PBC; perception of how convenient the behavior is and also said to influence behavior directly). Via these direct measures, intention is indirectly affected by behavioral beliefs (beliefs underlying attitudes reflecting a cost-benefit analysis of perceived advantages and disadvantages for performing the behavior), normative beliefs (beliefs underlying subjective norm representing the perceived approval of specific important referents) and control beliefs (beliefs underlying PBC that reflect factors that encourage or discourage performing the behavior).

Despite the debate about the utility of the TPB in health psychology applications (14), there is meta-analytic evidence in support of the TPB predictors (15-17) and the predictive validity of the TPB in healthy eating and physical activity has been demonstrated in many studies previously (18-24), including among samples of people diagnosed with type 1 and 2 diabetes (25-31). However, to design interventional studies, information regarding specific beliefs for behavioral decisions is necessary. Belief based assessments assist in the recognition of specific underlying beliefs, differentiating high intenders and low intenders that can then be used to inform targeted intervention strategies (32).

TPB belief-based studies for health-related behaviors such as activity and eating decisions have been undertaken previously (33-36) including examples for those people diagnosed with T2D (37). White et al., in their study among people diagnosed with T2D and cardiovascular disease, identified important beliefs differentiating performers and non-performers of physical activity included feeling healthy, feeling sore and laziness and important beliefs differentiating between those who followed low fat food consumption guidelines and those who did not including feeling healthy, using unfamiliar ingredients, and approval of family as well as friends and peers. In their study using a non-clinical sample, Armitage and Conner (1999) identified a number of behavioral beliefs regarding low-fat diet that differed between intenders and non-intenders including perception of a boring diet and feeling healthy (38). Intenders were also more likely to consider that they have more time and knowledge about consuming a low-fat diet. Although there are a number of TPB belief-based studies examining physical activity and low-fat food consumption including for at-risk populations like people diagnosed with T2D, there are no studies investigating the underlying beliefs for carbohydrate counting, often a key behavior undertaken and/or recommended for those with T2D. However, one study that has investigated food monitoring is Masula and Astrom’s (2003) study of sugar restriction among university students who found discrepancies such as intenders were more likely than non-intenders to perceive performing the health behavior to result in avoidance of overweight and preventing diabetes in old age (34). Further, non-intenders were more likely to consider having a boring diet as a negative belief. In the same research, all of the listed normative beliefs (e.g. approval of relatives, friends, etc.) were significant. Finally, feeling tired, feeling bored and having enough pocket money were the barrier control beliefs endorsed more highly by non-intenders.

Effective behavioral education entails the understanding of important beliefs that can affect intention and subsequently behavior. For this reason, the current study used the TPB theoretical framework to assess behavioral, normative and control beliefs regarding three critical behaviors of low-fat food consumption, carbohydrate counting and physical activity among an adult sample of people diagnosed with T2D. In an exploratory manner, we identified the important differences in beliefs differentiating between low and high intenders for each of the key target behaviors. Of the indirect measurements of behavioral, normative and control beliefs included in the study, due to limited time and space, the evaluative items were omitted given that the evaluative items are not essential for belief examination (11).

The current study adds to the literature in that it uses a theory-based belief comparison between high intenders and low intenders for key diabetes management behaviors including carbohydrate counting which has not been assessed previously in structured belief-based studies. Thus, the purpose of the present study was to investigate the relationship between belief-based measures and intention to perform three key diabetes management behaviors in adults diagnosed with T2D.

## Methods

### Participants

Ethical approval was received from the ethics committee of “Blinded for Review” University. Using a list of people diagnosed with diabetes provided by the “Blinded for Review”, individuals were selected using a computer-generated randomization pattern and then contacted to participate in the study. Those who agreed signed a written informed consent. A total of 115 adults with diabetes (aged over 25 years) consisting of 71 females (61.7%) and 44 males (38.3%) completed the main study. Demographic characteristics are shown in Table 1. The mean age of the participants was 48.7 years (SD=5.95; range=30-61 years). Most participants were full-time employees (n=61, 53%) and married (n=103, 90.4%). The mean number of years since diagnosis was 5.85 years (SD=2.89; range=2-14 years).

**Table 1.** Demographic characteristics of Participants.

| Variable |  | N=115 |  |
| --- | --- | --- | --- |
|  |  | Frequency | Percentage (%) |
| **Gender** |  |  |  |
|  | Male | 44 | 38.3 |
|  | Female | 71 | 61.7 |
| **Age** |  |  |  |
|  | ≤40 | 10 | 8.7 |
|  | 41-50 | 59 | 51.3 |
|  | ≥50 | 46 | 40 |
| **Educational Level** |  |  |  |
|  | Illiterate | 8 | 6.9 |
|  | Primary/Secondary School | 19 | 16.6 |
|  | High School | 13 | 11.4 |
|  | Diploma | 28 | 24.3 |
|  | Associate Degree | 22 | 19.1 |
|  | Bachelors' Degree | 23 | 20 |
|  | Masters' Degree | 2 | 1.7 |
| **Job Status** |  |  |  |
|  | Unemployed | 1 | 0.9 |
|  | Full-time | 61 | 53 |
|  | Part-time | 5 | 4.2 |
|  | Retired | 12 | 10.5 |
|  | Housekeeper | 36 | 31.4 |
| **Marital Status** |  |  |  |
|  | Married | 103 | 90.4 |
|  | Single | 4 | 3.5 |
|  | Divorced | 2 | 1.8 |
|  | Widow/Widower | 5 | 4.4 |

Participants in both an initial elicitation and main study were provided with definitions of the three target behaviors. The definition related to **low-fat food/ meal options** was reducing saturated fat intake by eating low-fat dairy products, using polyunsaturated and monounsaturated oils [plant based] and avoiding fried foods, trimming fat from meat [lean meat] (39, 40). The definition of **carbohydrate counting** was: identifying which foods contain carbohydrate, then assessing how much carbohydrate a serving of food (or an entire meal) contains and if you use insulin, match with insulin dose) (41, 42). The definition of **regular physical activity** was: engaging in moderate to vigorous physical activity for at least 150 minutes per week [half an hour/day, most days of the week]) (43, 44). In the main study, the definitions of the behaviors were repeated at intervals to remind participants of the target behaviors.

### Elicitation Study

To obtain salient behavioral, normative and control beliefs regarding the three target behaviors to include in the main questionnaire, an elicitation study was undertaken with 30 adults diagnosed with T2D using content analysis. A computer-generated pattern was used to obtain individuals in the elicitation phase who were representative of the main sample and didn’t engage in the main questionnaire phase. Their mean age was 44.93 years (SD=7.25; Range=29-58 years). Females comprised 60% (n=18) of the sample and males were 40% (n=12). The procedure for the elicitation study was based on the guidelines of Ajzen and Fishbein (10). Face-to-face individual interviews lasted for approximately 35 minutes per person. Open-ended questions asked participants to consider advantages and disadvantages of the three behaviors (for behavioral beliefs), important referents who may approve or disapprove of their performing the behaviors (normative beliefs), and facilitators and obstacles that may encourage or prevent performing the behaviors (control beliefs). Responses were content-analyzed, and the most commonly reported responses informed the belief-based measures in the main questionnaire.

The most frequent responses of elicitation study were revealed for low-fat food consumption (advantages; e.g. “reduces my blood lipids”/ disadvantages; e.g. “has a bad taste”, referents; e.g. “health care providers”, obstacles; e.g. “eating out in restaurants/ workplace or parties”, facilitators e.g. “knowledge of high-fat food effects or disease complications”), carbohydrate counting (advantages; e.g. “helps control my blood sugar/ lowers my blood sugar”/ disadvantages; e.g. “it’s too boring”, referents; e.g. “health care providers”, obstacles; e.g. “laziness”, facilitators e.g. “knowledge of carbohydrate counting” and physical activity (advantages; e.g. “helps control my weight/ my weight loss”/ disadvantages; e.g. “makes me tired”, referents; e.g. “spouse”, obstacles; e.g. “lack of time”, facilitators e.g. “others exercising with me”) and these were used in the main questionnaire. It should also be noted that for physical activity, two additional disadvantages (“puts my health at risk” and “may result in injury”) were added following a literature review of beliefs shown to influence people’s activity-related behaviors.

### Main Questionnaire

#### Belief measures

The participants rated how likely it would be that the advantages and disadvantages obtained from the elicitation study would occur if they performed each of the target behaviors (behavioral beliefs). Participants also rated how likely the important referents obtained from the elicitation study would think that they should perform the behaviors (normative beliefs). The control beliefs items asked participants to rate how likely it was that the facilitators and barriers obtained from the elicitation study would encourage and prevent them, respectively, in relation to engaging in the target behaviors. All measures were assessed on a 7-point Likert scale (from 1 **extremely unlikely** to 7 **extremely likely**, and, where appropriate, included the option of “**doesn’t apply to me**”).

#### Intention

Intention for each of the three behaviors was assessed by two items, “I intend to **consume low-fat foods**/ **use carbohydrate counting method**/ **engage in regular physical activity** over the next month” and “It is likely that I will **consume low-fat food**/ **use carbohydrate counting method**/ **engage in regular physical activity** over the next month”. The two-items for each behavior were based on a 7-point Likert scale (strongly disagree (1) to strongly agree (7)) averaged to produce an intention scale. Pearson correlations for the low-fat food, carbohydrate counting and physical activity intentions scales were r(115)=0.82, p<0.001, r(113)=0.73, p<0.001 and r(113)=0.65, p<0.001, respectively.

## Results

All data were analyzed using SPSS (version 22: IBM corporation). The mean scores for the intention scales for low-fat foods, carbohydrate counting and physical activity were 5.73 (SD=0.81), 5.70 (SD=0.89) and 5.64 (SD=0.82), respectively. For all three behaviors, these intentions are considered to be fairly high. Three multivariate one-way analysis of variance (MANOVAs) were performed for investigating behavioral, normative and control belief differences between low intenders and high intenders, where beliefs were entered as the dependent variables and intention to perform the behavior as the independent variable. Across all three behaviors those who rated their intention at or above the scale mid-point (“4”) were considered as high intenders and those who rated below “4” were considered as low intenders.

### MANOVA: Low-fat food

There were no significant multivariate effects (using Wilk’s criterion) found for behavioral F(1,98)=1.714, p=0.078, normative F(1,43)=0.438, p=0.848, and control F(1,99)=1.139, p=0.344, beliefs for low-fat food consumption. Table 2 provides the univariate results for the beliefs about low-fat food choice.

**Table 2.** Mean Behavioral, Normative and Control Beliefs Among Low intenders and High intenders Regarding Low-fat Food Option.

| Low-fat food option | Low intenders | High intenders |
| --- | --- | --- |
|  | Mean | Mean |
| **Behavioral Beliefs** | N=56 | N=55 |
| Help control my weight/ my weight loss | 5.75 | 6.05 |
| Make me healthier | 5.89 | 6.29 |
| Make me feel good/ feel light | 5.33 | 5.81 |
| Reduce my blood lipids | 5.91 | 6.29 |
| Lower my blood pressure | 4.41 | 4.90 |
| Reduce/ help control my blood sugar levels | 4.32 | 4.92 |
| Prevent diabetic complications | 5.50 | 5.78 |
| Lead to constipation | 4.25 | 3.89 |
| Have a bad taste | 4.75 | 4.27 |
| Make it harder for me to prepare meals | 3.50 | 2.90 |
| Make me worry about too much loss of blood lipids | 1.87 | 2.05 |
| Lead to long-term complications | 1.89 | 2.16 |
| **Normative Beliefs** | N= 57 | N=57 |
| Spouse | 6.10 | 5.92 |
| Children | 5.43 | 5.68 |
| Other family members | 4.91 | 4.71 |
| Friends & acquaintances | 4.84 | 4.80 |
| Colleagues | 5.92 | 5.91 |
| Health care provider | 6.05 | 6.05 |
| **Control Beliefs** | N=55 | N=55 |
| Lack of food variety | 3.76 | 4.10 |
| Eating out in restaurants/ workplace or parties* | 5.30 | 4.70 |
| Laziness | 4.10 | 4.23 |
| Lack of time | 2.89 | 3.01 |
| Lack of knowledge | 5.72 | 5.45 |
| Family's preparation of high fat foods | 4.83 | 4.87 |
| Knowledge of high-fat-food effects/ disease complications | 6.09 | 6.21 |
| Media advertisement/ books & magazines providing helpful information | 4.85 | 5.03 |
| Medical advice from health professionals | 5.90 | 6.18 |

Using Wilk’s criterion, there was a significant multivariate effect of behavioral beliefs for carbohydrate counting, F(1,112)=3.548, p=0.001. Univariate tests (Table 3) revealed differences between high intenders and low intenders where high intenders considered weight control F(1,112)=4.207, p=0.043, making them healthier F(1,112)=20.988, p<0.001, feeling good F(1,112)=4.539, p=0.035, and controlling diabetic complications F(1,112)=5.561, p=0.020 as more likely outcomes of carbohydrate counting than low intenders. High intenders were also less likely than low intenders to believe that difficulty of food preparation F(1,112)=6.464, p=0.012, boringness F(1,112)=11.550, p=0.001, food restriction F(1,112)=4.387, p=0.039, and taking too much attention F(1,112)=4.397, p=0.038, would be outcomes of carbohydrate counting.

**Table 3.** Mean Behavioral, Normative and Control Beliefs Among Low intenders and High intenders Performing Carbohydrate Counting

| Carbohydrate counting | Low intenders | High intenders |
| --- | --- | --- |
|  | Mean | Mean |
| **Behavioral Beliefs** | N=44 | N=68 |
| Help control my weight/ my weight loss* | 4.70 | 5.27 |
| Make me healthier** | 5.36 | 6.25 |
| Reduce/ help control my blood sugar levels | 5.81 | 6.13 |
| Make me feel good/ feel light* | 4.86 | 5.44 |
| Prevent diabetic complications* | 5.50 | 6.01 |
| Make it harder for me to prepare meals* | 4.15 | 3.35 |
| Make me tired because it's too boring* | 4.75 | 3.55 |
| Restrict foods I can eat* | 5.68 | 5.04 |
| Take too much of my attention* | 5.29 | 4.67 |
| **Normative Beliefs** | N=43 | N=69 |
| Spouse* | 5.39 | 6.01 |
| Children | 5.79 | 5.84 |
| Other family members | 5.04 | 4.63 |
| Friends & acquaintances | 4.39 | 4.72 |
| Colleagues | 5.67 | 5.86 |
| Health care provider | 6.02 | 6.24 |
| **Control Beliefs** | N=44 | N=68 |
| Laziness | 5.00 | 4.69 |
| Eating out in restaurants/ workplace or parties | 5.47 | 5.38 |
| Hunger * | 4.86 | 5.58 |
| Lack of willpower | 4.20 | 3.98 |
| Lack of time | 3.15 | 3.50 |
| Lack of knowledge about how to count carbohydrates | 5.77 | 5.98 |
| Medical advice from health professionals** | 5.81 | 6.25 |
| Knowledge of carbohydrate counting** | 5.70 | 6.33 |
| Media advertisement/ books & magazines providing helpful information | 4.15 | 4.36 |
| **is significant at p<0.01 |  |  |
| *is significant at p<0.05 |  |  |

There was a significant multivariate effect of normative beliefs for carbohydrate counting, F(1,112)=2.894, p=0.012. The results of univariate analyses indicated that high intenders were more likely than low intenders to perceive their spouse F(1,112)=4.554, p=0.035, as approving of their carbohydrate counting behavior.

There was also a significant multivariate effect of control beliefs for carbohydrate counting, F(1,112)=3.966, p<0.001. Unexpectedly, high intenders were more likely to perceive “hunger” F(1,112)=6.438, p=0.013, as a barrier to carbohydrate counting than low intenders. High intenders were more likely than low intenders to consider medical advice F(1,112)=9.241, p=0.003, and the knowledge about how to do carbohydrate counting F(1,112)=17.777, p<0.001, as likely facilitators of behavioral performance.

### MANOVA: Physical activity

MANOVA results revealed a significant multivariate effect of behavioral beliefs for physical activity, F(1,110)=2.810, p=0.004. Univariate analyses indicated that high intenders were more likely than low intenders to consider that feeling good F(1,110)=10.902, p=0.001, reducing/ helping control blood sugar F(1,110)=13.927, p<0.001, and making themselves healthier F(1,110)=4.840, p=0.030, were likely outcomes of performing the behavior. Low intenders were more likely than high intenders to perceive that tiredness F(1,110)=14.787, p<0.001 would be an outcome.

The multivariate effect was not significant for normative beliefs F(1,113)= 0.790, p=0.579, but was significant for control beliefs, F(1,109)= 2.385, p=0.014. Univariate results showed a significant difference in the cost of exercise F(1,109)=16.974, p<0.001, whereby high intenders perceived high cost as a less likely barrier to stop engaging in regular physical activity during the next month than low intenders. Further, low intenders considered media advertisement, books and magazines F(1,109)=5.599, p=0.020, as less likely facilitators for physical activity than high intenders (Table 4).

**Table 4.**
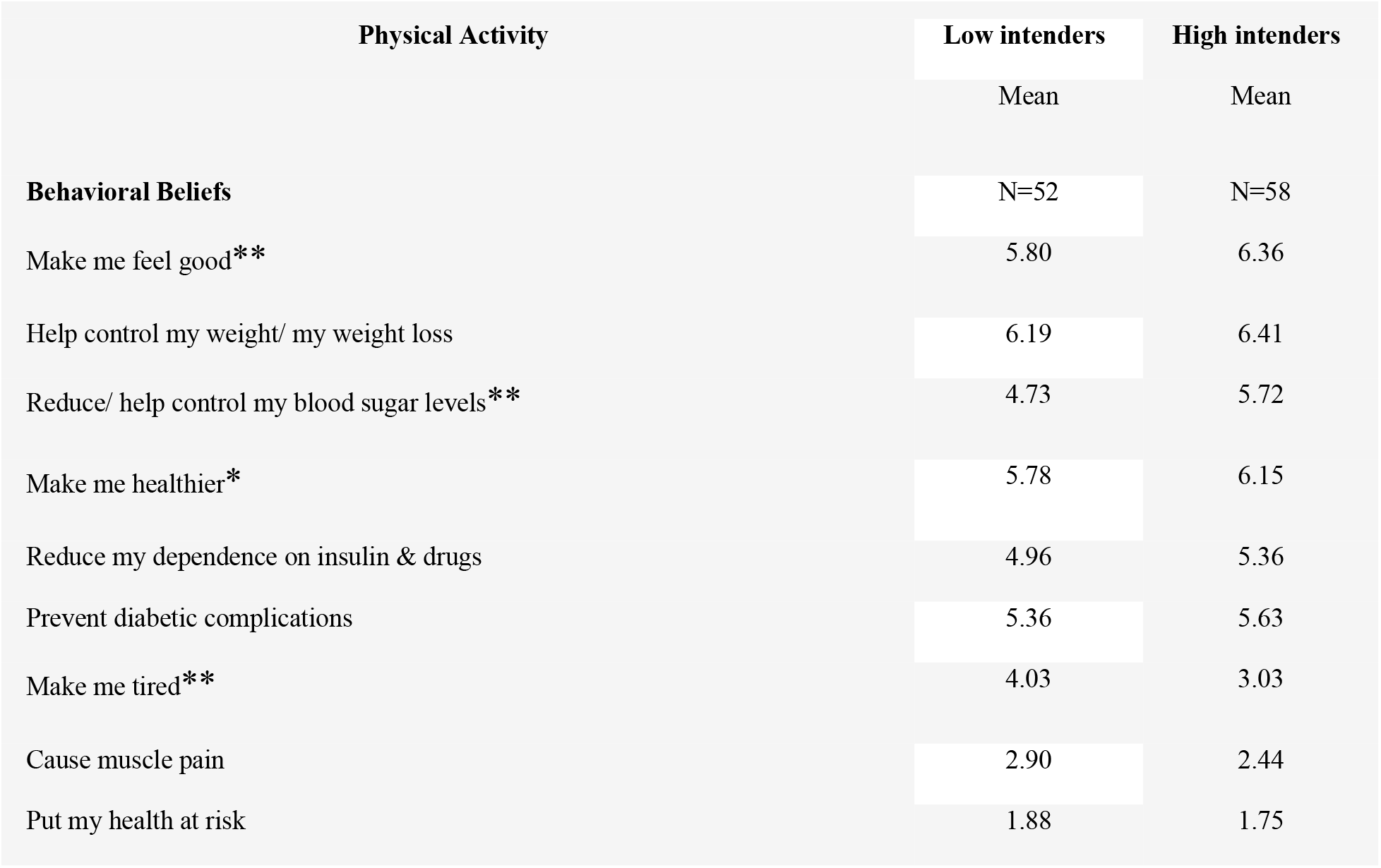
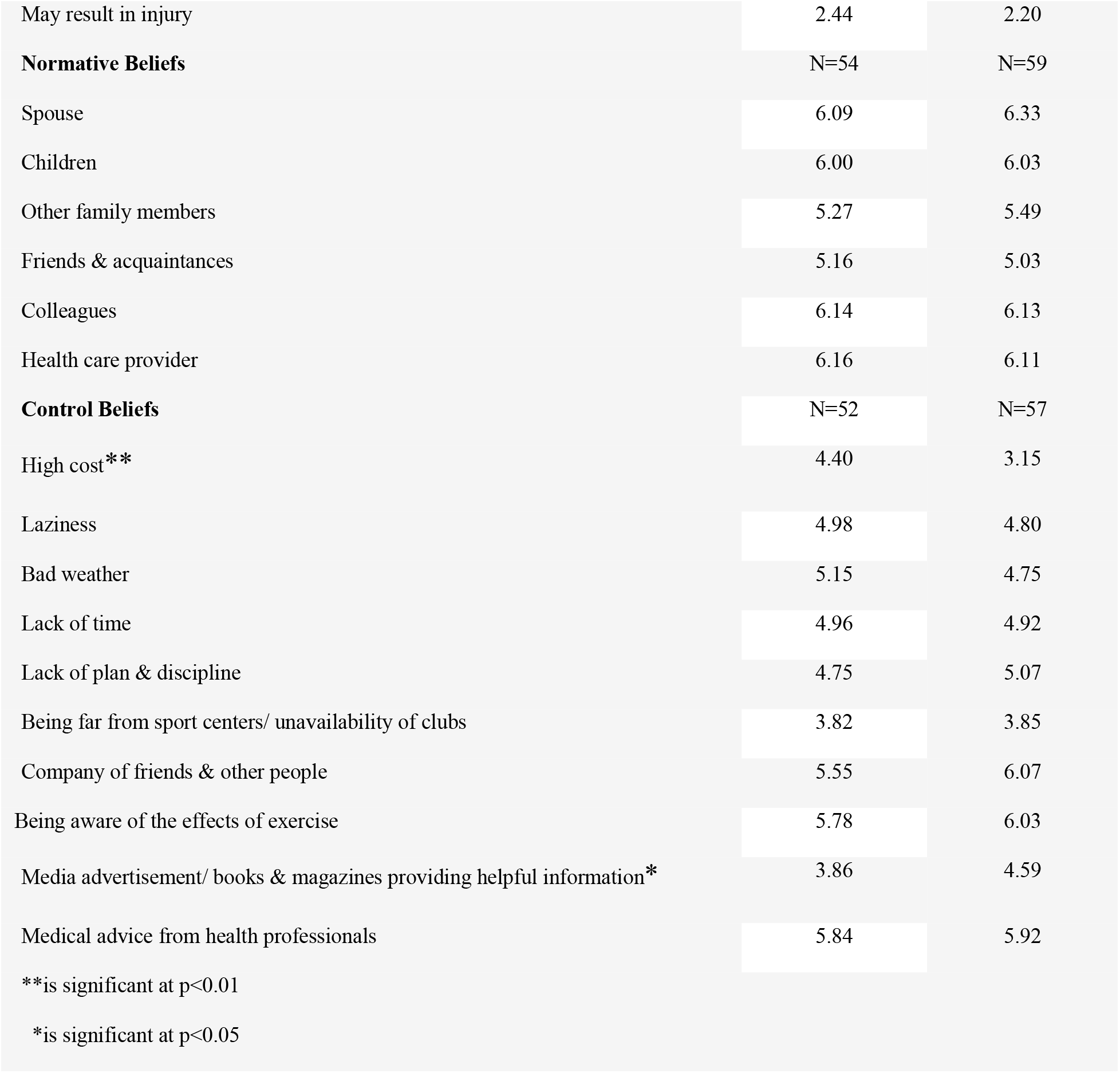
Mean Behavioral, Normative and Control Beliefs Among Low intenders and High intenders Engaging in Physical Activity.

| Physical Activity | Low intenders | High intenders |
| --- | --- | --- |
|  | Mean | Mean |
| **Behavioral Beliefs** | N=52 | N=58 |
| Make me feel good** | 5.80 | 6.36 |
| Help control my weight/ my weight loss | 6.19 | 6.41 |
| Reduce/ help control my blood sugar levels** | 4.73 | 5.72 |
| Make me healthier* | 5.78 | 6.15 |
| Reduce my dependence on insulin & drugs | 4.96 | 5.36 |
| Prevent diabetic complications | 5.36 | 5.63 |
| Make me tired** | 4.03 | 3.03 |
| Cause muscle pain | 2.90 | 2.44 |
| Put my health at risk | 1.88 | 1.75 |
| May result in injury | 2.44 | 2.20 |
| **Normative Beliefs** | N=54 | N=59 |
| Spouse | 6.09 | 6.33 |
| Children | 6.00 | 6.03 |
| Other family members | 5.27 | 5.49 |
| Friends & acquaintances | 5.16 | 5.03 |
| Colleagues | 6.14 | 6.13 |
| Health care provider | 6.16 | 6.11 |
| **Control Beliefs** | N=52 | N=57 |
| High cost** | 4.40 | 3.15 |
| Laziness | 4.98 | 4.80 |
| Bad weather | 5.15 | 4.75 |
| Lack of time | 4.96 | 4.92 |
| Lack of plan & discipline | 4.75 | 5.07 |
| Being far from sport centers/ unavailability of clubs | 3.82 | 3.85 |
| Company of friends & other people | 5.55 | 6.07 |
| Being aware of the effects of exercise | 5.78 | 6.03 |
| Media advertisement/ books & magazines providing helpful information* | 3.86 | 4.59 |
| Medical advice from health professionals | 5.84 | 5.92 |
| **is significant at $p<0.01$ | | |
| *is significant at $p<0.05$ | | |

## Discussion

We examined the behavioral, normative and control beliefs for three key health behaviors (low-fat food consumption, carbohydrate counting and physical activity) among adults diagnosed with T2D. A range of beliefs differentiated between low- and high intenders that can be used to inform intervention studies aimed at improving diabetes management for those diagnosed with this chronic illness.

For low-fat food consumption, unexpectedly, no significant effects emerged. This finding might be due to the fact that diabetes is so closely aligned in people’s minds with changing their diet to lower high fat foods that the groups did not differ substantially on beliefs. However, inspection of the means for the beliefs for low-fat food consumption did not appear substantially higher than for the other two management behaviors.

The results did, however, reveal significant discrepancies in behavioral beliefs among low and high intenders for carbohydrate counting which suggest that highlighting beliefs such as weight control, improving one’s health, feeling good and controlling diabetic complications may be the best targets for intervention. It seems high intenders are more concerned with carbohydrate counting and have broader knowledge about it, since weight control and diabetic complications especially in the long term are related to carbohydrate counting (45-47). Thus, interventions designed to promote carbohydrate counting in adults with T2D should focus on the health influences of weight loss (e.g., improving cardiovascular health) and metabolic outcomes (i.e., reducing glycaemia, blood pressure and improving lipid profile, as well as decreasing mortality rates (48, 49)). Importantly, in relation to the importance of weight loss in diabetes, it has been reported that even intention to lose weight, without actually leading to weight loss, can improve outcomes probably due to healthy behaviors accompanied by the attempt to lose weight (49). In addition to strategies targeting the likelihood of positive health-associated feelings, the identified disadvantages of difficulty of food preparation, food restriction and focusing too much attention should be challenged or their impact diminished. Hence, interventions should incorporate strategies to facilitate food preparation methods by educating individuals how to incorporate tasty foods that lack carbohydrates (e.g. lean meats, fish, eggs, etc.) or contain less carbohydrates (non-starchy vegetables such as lettuce, spinach, celery, parsley, etc.). Additionally, health care providers should explain that food elimination is not a necessary strategy but instead aim to distribute carbohydrate foods evenly in meals and snacks. Further, intervention and educational efforts should focus on planning strategies that facilitate managing carbohydrate counting so that it is not perceived as burdensome.

Among normative beliefs, intenders rather than non-intenders viewed their spouse as a social referent more likely to persuade them to engage in carbohydrate counting. Therefore, future interventions should encourage spouses to participate in education sessions as supporters of healthy behavior decisions or encourage them to be a part of a formal commitment to implement changes (e.g., co-signing a healthy behavior ‘contract’).

Unexpectedly, hunger was regarded as a barrier more by high intenders than low intenders. Slow paced eating has been shown to reduce hunger sensation in overweight and obese people with T2D (50); thus, interventions that educate slow paced eating may be beneficial for those people committed to trying carbohydrate counting but who are concerned about hunger pangs. Further, encouraging self-control and incorporation of non-carbohydrate or low-carbohydrate foods with the emphasis to consume in moderation can induce fullness sensation. On the basis of the findings, behavioral interventions also should improve the knowledge of carbohydrate counting for people diagnosed with diabetes by holding instructional classes with practical guidance included. It is also suggested to involve medical professionals in advice and encouragement as high intenders perceive approval from this referent as more likely to facilitate behavioral performance.

For physical activity, the results indicate that interventional approaches should promote perceptions of feeling good, controlling blood sugar and improved health as positive outcomes. Consistent with our findings, White et al., in their study in older adults with T2D and CVD found feeling healthy as a belief differentiating performers and non-performers of physical activity (37). Tiredness associated with engaging in physical exercise as a negative outcome could be managed by motivating people diagnosed with T2D to engage in a range of interesting activities, with rest periods, to combat fatigue. Costs was one of the control beliefs which differed between high- and low intenders. Therefore, it is recommended that programs encourage low-cost activities such as neighborhood walks.

By examining the three key behaviors simultaneously, this study produces a triad of critical behaviors for diabetes management. In addition to the small sample size, one main limitation of the research is the cross-sectional design and future studies should also employ a prospective design to measure performance of the target behaviors. The study included “Blinded for Review” members who may have been highly motivated, bringing into question the generalizability of the results to the majority of people with diabetes unconnected to the “Blinded for Review”. With respect to the growing T2D epidemic in adolescents and children, it is recommended future studies investigate important beliefs for these three behaviors among these age groups to assist in the design of prevention programs. Furthermore, the importance of carbohydrate counting is supported for individuals diagnosed with type 1 diabetes (51-54), a gap in belief assessment that warrants future research.

Overall, the present study identified the factors distinguishing between low and high intenders for the belief base of the TPB in the context of diet-related and physical activity health issues in a population diagnosed with T2D. Identifying the important underlying beliefs related to people’s intentions adds to the extant literature and enables the development of practical recommendations for use in resultant health interventions, an important component in efforts to reduce the burden of this chronic health condition on both those affected and broader society.

## Data Availability

Data cannot be shared publicly because of ethical mandates. Data are available from the Ethics Committee of the Tehran University of Medical Sciences (please contact Reza Daryabeygi and he will contact the committee on behalf) for researchers who meet the criteria for access to confidential data.

## Acknowledgment

This research was supported by vice chancellor for research, Tehran University of Medical Sciences, Iran. In the memory of Professor Hossein Ghasemi (deceased) who illuminated the idea of the current behavioural study and special thanks to Dr. Louise Starfelt for her generous assists on the statistical analysis. We would also like to thank Dr. Soheil Hassanipour Azgomi, Mr. Iliyar Yamrali, Mr. Reza Ghadiri Rad, Mr. Rezaei, Ms. Bani and Ms. Fouladvand for assistance with data collection.

